# Evaluation of the Liberty16 mobile real time PCR device for use with the SalivaDirect assay for SARS-CoV-2 testing

**DOI:** 10.1101/2021.08.08.21261746

**Authors:** Devyn Yolda-Carr, Darani A. Thammavongsa, Noel Vega, Susan J. Turner, Paul J. Pickering, Anne L. Wyllie

## Abstract

The COVID-19 pandemic has highlighted the need and benefits for all communities to be permitted timely access to on-demand screening for infectious respiratory diseases. This can be achieved with simplified testing approaches and affordable access to core resources. While RT-qPCR-based tests remain the gold standard for SARS-CoV-2 detection due to their high sensitivity, implementation of testing requires high upfront costs to obtain the necessary instrumentation. This is particularly restrictive in low-resource settings. The Ubiquitome Liberty16 system was developed as an inexpensive, portable, battery-operated single-channel RT-qPCR device with an associated iPhone app to simplify assay set-up and data reporting. When coupled with the SalivaDirect protocol for testing saliva samples for SARS-CoV-2, the Liberty16 device yielded a limit of detection (LOD) of 12 SARS-CoV-2 RNA copies/µL, comparable to the upper end of the LOD range for the standard SalivaDirect protocol when performed on larger RT-qPCR instruments. While further optimization may deliver even greater sensitivity and assay speed, findings from this study indicate that small portable devices such as the Liberty16 can deliver reliable results and provide the opportunity to further increase access to gold standard SARS-CoV-2 testing.

## Introduction

Timely access to SARS-CoV-2 tests remains a crucial factor in effective clinical and community-wide management of COVID-19^1^. To date, RT-qPCR-based tests remain the gold standard due to their high sensitivity, yet implementation of testing requires high upfront costs to obtain the necessary instrumentation which is particularly restrictive in low resource settings. To overcome the limitations of traditional RT-PCR testing, the Liberty16 system (Ubiquitome Ltd, Auckland, New Zealand) was developed as an inexpensive, portable, battery-operated single-channel RT-qPCR device with an associated iPhone app to simplify assay set-up and data reporting. Similarly, the SalivaDirect PCR-based SARS-CoV-2 assay^2^ was developed to expand testing capacity by removing specialized reagents, equipment, and time components to decrease the time and cost of nucleic acid extraction. As the SalivaDirect protocol can still be constrained in resource-limited settings by lack of access to RT-PCR devices, we evaluated the performance of the novel Liberty16 system for detection of SARS-CoV-2 in saliva using a singleplex version of the SalivaDirect workflow.

## Methods

### Limit of Detection

A limit of detection (LOD) range finding study was conducted to compare a single-plex version of the SalivaDirect protocol^3^ run on the Liberty16 System as compared to the standard dualplex protocol run on the BioRad CFX96 Touch. The Liberty16-modified protocol used the CDCs FAM-labelled N1 and RNaseP (RP) primer probe sets, tested on saliva lysates in separate reactions. Primer and probe concentrations were the same as that used for the SalivaDirect assay. In all other respects the assay was performed as described in the SalivaDirect protocol using the Thermo Fisher Proteinase K (A42363) and New England Biolabs Luna Universal Probe One-Step RT-qPCR (2x) kit (E3006).

For the LOD range finding study, a SARS-CoV-2 positive saliva specimen, collected in accordance with the Yale University HIC-approved protocol #2000027690, with a known virus concentration (3.7 × 104 copies/µL) was spiked into saliva negative (HIC-approved protocol #2000027690) for SARS-CoV-2 using the CDC assay^4^. The following 2-fold dilution series was tested in triplicate to determine the preliminary LOD: 100, 50, 25, 12.5, 6.25, 3.125, and 1.5 copies/µL. The preliminary LOD was then confirmed with 20 additional replicates.

### Confirmatory testing and protocol optimization

For the testing of clinical specimens 31 de-identified saliva specimens previously tested by SalivaDirect were selected for assay verification (Yale University HIC-approved protocol #2000029551). The 31 specimens represented an array of 30 positive samples (Ct values 23-39.8) and 1 negative sample (Ct value ND, not detected). The saliva samples were processed by the SalivaDirect protocol, with PCR-testing performed on the Liberty16 device. Each sample was tested for SARS-CoV-2 N1 twice using both the standard SalivaDirect PCR settings (95°C, 10s; 55°C, 30s) and an additional fast cycling protocol (95°C, 2s; 55°C, 5s). The same reverse transcriptase activation conditions (52°C, 10min; 95°C, 2 min) were used for both runs. Differences between the two protocols were assessed for statistical significance using the Wilcoxon Signed Rank test for non-parametric data.

## Results

### The LOD of the Liberty16 SalivaDirect assay is comparable to other PCR instruments

A preliminary range-finding study using triplicate samples across a two-fold dilution series indicated an LOD of between 6 copies/µL (0/3 samples detected) and 12 copies/µL (3/3 detected; mean Ct value = 37.7). The same samples run on the Biorad CFX96 Touch using the standard dualplex assay at 12 copies/µL yielded an average Ct value of 36.00. The LOD for the Liberty16 system was confirmed at 12 copies/µL with 20/20 samples positive and an average Ct value of 35.18 (standard deviation = 0.71; Figure 1).

**Figure 1.**
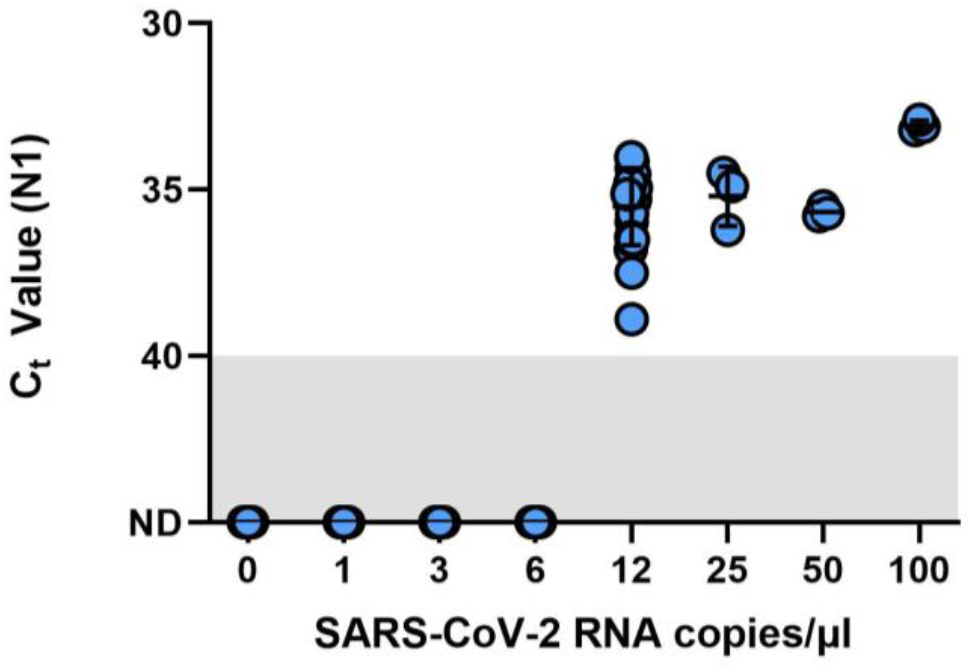
Limit of detection (LOD) finding study for the SalivaDirect protocol adapted for RT-PCR testing by the Liberty16 instrument. A SARS-CoV-2 positive saliva specimen with a known virus concentration (3.7 × 104 copies/µL) was spiked into saliva negative for SARS-CoV-2 to create the following 2-fold dilution series: 100, 50, 25, 12.5, 6.25, 3.125, and 1.5 copies/µL. Spiked saliva samples were tested in triplicate to determine the preliminary LOD using the SalivaDirect protocol and tested using the NEB Luna 2x PCR mastermix and run on the Liberty16 system using the standard PCR (95°C, 10s; 55°C, 30s) and reverse transcriptase activation conditions (52°C, 10min; 95°C, 2 min). The preliminary LOD was determined as between 6 copies/µL (0/3 samples detected) and 12 copies/µL (3/3 detected). An additional 20 replicates of 6 copies/µL and 12 copies/µL were tested, confirming the LOD for the Liberty16 system as 12 copies/µL with 20/20 samples positive.

### Fast cycling protocol reduces run time without compromising N1 detection

As compared to the standard SalivaDirect protocol, the fast PCR cycling protocol completed in under one hour, saving more than 20 minutes. Despite the fast run time, comparative analysis of clinical samples (30 positive and 1 negative) revealed a high level of concordance between N1 values for the fast and regular protocols (median Ct difference = 1.37; Wilcoxon *p* > 0.1; Figure 2). Ten samples were not detected (NA) by either protocol. All were previously shown to have Ct value at or below the limit of detection of the Liberty16 device running the SalivaDirect assay.

**Figure 2.**
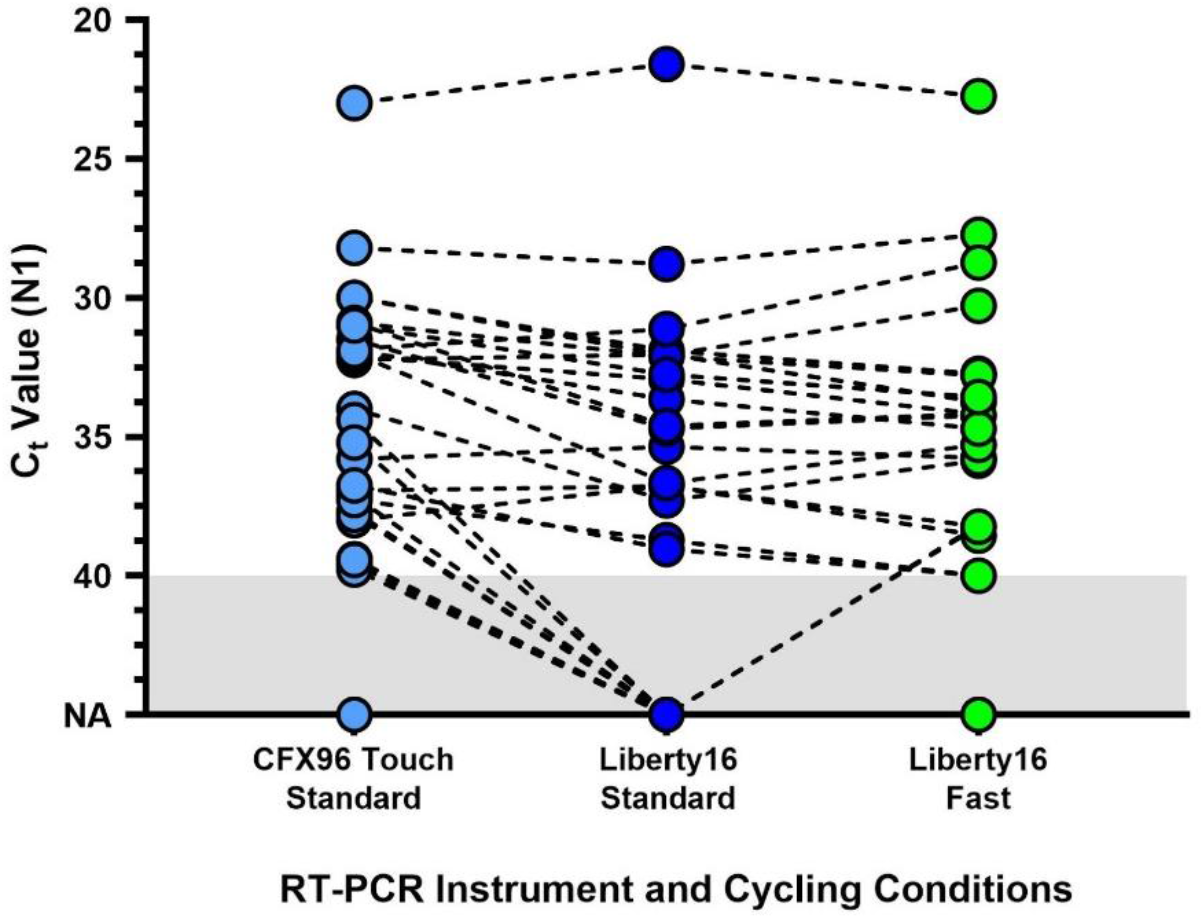
Comparison of the standard SalivaDirect PCR and updated fast cycling protocols on SARS-CoV-2 virus RNA N1 detection. De-identified clinical saliva samples, previously tested using the standard SalivaDirect protocol on the CFX96 Touch, using the NEB Luna 2x PCR mastermix were also run on the Liberty16 system using the standard RT-qPCR (95°C, 10s; 55°C, 30s) or fast RT-qPCR (95°C, 2s; 55°C, 5s) cycling conditions. The same reverse transcriptase activation conditions (52°C, 10min; 95°C, 2 min) were used for both runs. The resulting SARS-CoV-2 N1 (Ct) values did not differ between either Liberty16 protocol (Wilcoxon p > 0.1). Two out of 21 sample pairs (10%) yielded not detected (ND) values when run using the standard protocol, while 2 pairs yielded Ct values of 40 and 41 respectively. Ten samples, previously positive when tested on the CFX96 Touch were not detected by either Liberty16 protocol. All were previously shown to have Ct values at or below the limit of detection of the Liberty16 device running the SalivaDirect assay.

With the current throughput of samples using the SalivaDirect protocol on the Liberty16 device being 6 samples plus two controls per ∼80-minute run, this would allow for 5 complete runs (30 samples) per day. The faster run time enables completion of 8 runs per day, increasing the sample throughput by 60% to 48 samples per day.

## Conclusions

Access to reliable SARS-CoV-2 testing and to vaccines are the two factors currently dividing humanity in the race to minimise COVID-19 mortality and long-term morbidity. Until global vaccination rates reach levels sufficient to provide herd immunity, the first line of defence against widespread disease is timely and accurate screening for infection across all sectors of the community^5^. Current barriers to testing include public dislike of inconvenient and/or uncomfortable sample collection means such as clinician-administered nasopharyngeal swabs as well as availability of specialist testing equipment, reagents and expertise for sample analysis^6^. Use of saliva as the testing matrix resolves the first issue, being non-invasive and able to be carried out with simple instructions by a non-specialist in the home environment^7^.

The SalivaDirect protocol offers a range of RT-qPCR device and reagent options with reported LODs ranging from 3-12 copies/µL^8^ with the singleplex protocol performed on the Liberty16 device yielding results comparable to the upper end of that LOD range. While the throughput of the Liberty16 device is modest (6 samples/run) its portability, and comparatively low cost (USD $5,995), make this an affordable option for standing up new testing capability in remote and resource constrained environments. Furthermore, the initial run-time optimization reported here signals an opportunity to increase sample throughput by more than 60% by simply modifying the PCR run protocol. While further optimization may deliver even greater sensitivity and assay speed, this initial study indicates that small portable devices such as the Liberty16 can deliver reliable results and provide the opportunity to further increase access to gold standard testing capability.

Looking to the future, the COVID-19 pandemic has highlighted the need and benefits for all communities to be permitted timely access to on-demand screening for infectious respiratory diseases, including seasonal viruses such as RSV and influenza. This can be achieved with simplified testing approaches and affordable access to core resources.

## Data Availability

All raw data is available from the corresponding author.

## Author contributions

S.J.T., P.J.P and A.L.W. conceived the study. D.Y-C., S.J.T. and A.L.W. developed the study protocol. D.Y-C., D.A.T. and N.V. performed the experimental work. D.Y-C, S.J.T. and A.L.W. analyzed the data. D.Y-C., S.J.T. and A.L.W. wrote and edited the manuscript. All other co-authors reviewed, edited and approved the manuscript.

## Acknowledgements

This work was partially funded by a Fast Grant from Emergent Ventures at the Mercatus Center at George Mason University (A.L.W).

